# CLINICAL PERFORMANCE OF THE CALL SCORE FOR THE PREDICTION OF ADMISSION TO ICU AND DEATH IN HOSPITALIZED PATIENTS WITH COVID-19 PNEUMONIA IN A REFERENCE HOSPITAL IN PERU

**DOI:** 10.1101/2021.02.09.21250884

**Authors:** Rafael Pichardo-Rodriguez, Marcos Saavedra-Velasco, Willy Peña-Oscuvilca, Jhonnathan Ascarza-Saldaña, Cesar Sanchez-Alvarez, Gino Patron-Ordoñez, Oscar Ruiz-Franco, Jhony A. De La Cruz-Vargas, Herney Andres Garcia-Perdomo

## Abstract

**Objective:** Determine the CALL SCORE’s diagnostic accuracy for the prediction of ICU admission and death in patients hospitalized for COVID-19 pneumonia in a reference hospital in Peru.

**Methods:** We performed an analytical cross-sectional observational study. We included patients with COVID-19 pneumonia treated at the “Dos de Mayo” National Hospital. Patients over 18 years old with a diagnosis confirmed by rapid or molecular testing were included. Those with an incomplete, illegible, or missing medical history and/or bacterial or fungal pneumonia were excluded. Data were extracted from medical records. The primary outcomes were mortality and admission to the ICU. The Call Score was calculated for each patient (4 to 13 points) and classified into three risk groups. Summary measures were presented for qualitative and quantitative variables. The area under the model curve and the operational characteristics (sensitivity, specificity) were calculated for the best cut-off point.

**Results:** The Call Score reported an area under the curve of 0.59 (IC95%: 0.3 to 0.07), p = 0.43 for predicting death. However, for a cut-off point of 5.5, a sensitivity of 87%and a specificity of 65%were obtained. The area under the curve for ICU admission was 0.67 (95%CI: 0.3 to 0.07), p = 0.43; the 5.5 cut-off point showed a sensitivity of 82%and a specificity of 51%.

**Conclusions:** The Call Score shows a low performance for predicting mortality and admission to the ICU in Peruvian patients.

## INTRODUCTION

The new coronavirus disease 2019 (COVID-2019) has generated a global health crisis that has placed international public health in great danger[1]. Worldwide, 44 358 000 confirmed cases are reported, of which approximately 1 174 624 people have died from the disease[2]. Peru is no stranger to this problem; currently, approximately 891,000 new cases have been estimated in the country, with a total of 34,197 deaths[2]. To date, the SARS-CoV-2 infection continues to spread, and newly infected people grow every day, despite the isolation and quarantine measures established worldwide.

As the pandemic grows, we acknowledge that comorbidities such as obesity, arterial hypertension, and type 2 diabetes mellitus are risk factors and essential predictors for severity and death in patients with COVID-19 pneumonia[3,4]. Based on those mentioned above and due to the need for a more practical, rapid, and valid approach, different clinical prediction rules have been developed. However, most are based on complex calculations with measurements of variables that we do not have in our country [5,6].

The CALL SCORE was developed in China, a clinical tool based on four variables that facilitate patient decision-making with COVID-19 to achieve a practical and valid clinical prediction tool[3]. However, it is not validated for Latin American populations; its performance has only been evaluated in the Italian population with important results. It is crucial to evaluate its predictive impact and clinical performance in real clinical conditions on important outcomes such as mortality and ICU admission. Both, due to the lack of resources in our society and the need to improve our patients’ management and make an adequate triage of those who require specialized treatment[3].

The objective was to determine the CALL SCORE’s diagnostic accuracy for predicting admission to ICU and death in patients hospitalized due to COVID-19 pneumonia in a reference hospital in Peru.

## METHODS

### Study Design

We conducted a retrospective analytical cross-sectional study based on the guidelines of the STROBE[7] statement. The study was registered in the PRISA database for observational studies of the National Health Institute of Peru, with the following registration code: EI00000001429 (https://prisa.ins.gob.pe/index.php/acerca-de-prisa/busqueda-de-proyectos-de-investigacion-en-salud/1264-rendimiento-clinico-del-call-score-para-la-prediccion-de-ingreso-a-uci-y-muerte-en-pacientes-hospitalizados-con-neumonia-por-covid-19-en-un-hospital-de-referencia-en-peru). The study was evaluated and approved by the Office of Support for Teaching and Research of the “Dos de Mayo” National Hospital with registration number 027660-2020.

### Context

It was carried out in patients diagnosed with COVID-19 pneumonia treated at the “Dos de Mayo” National Hospital between April to July 2020.

### Participants

Patients older than 18 years with a confirmed diagnosis of COVID-19 by rapid or molecular testing were included, and those with an incomplete, illegible or missing medical history and/or bacterial or fungal pneumonia were excluded. The patients were registered in the COVID-19 hospital wards registry book, used as a source for the sampling frame and sample selection. The patients were selected using a simple random probability sampling process.

### Variables

Death was defined as the passing of the patient during his/her hospital stay. Admission to the ICU was defined as the patient’s admission to the ICU while hospitalized in COVID-19 wards. The CALL SCORE was calculated with de data taken at the admission for each patient spanning a range of 4 to 13 points (comorbidity 1-4 points, age 1-3 points, lymphocytes 1-3 points, LDH 1-3 points). Additional variables such as sex, age, comorbidities, serum lactate dehydrogenase (LDH), and absolute lymphocytes were included to describe the patients’ characteristics.

### Data Source

The data were extracted from the medical records of the selected patients and recorded in a standardized instrument developed based on the study’s objectives.

### Sample size

We required a minimum of 51 patients (Power 80%, an estimated area under curve 0.76, and a distribution ratio of 4.6) 6. It was calculated on the page’s web application: http://www.biosoft.hacettepe.edu.tr/easyROC/).

### Quantitative variables

The CALL SCORE classifies into three risk groups according to their probabilities of progression of COVID-19 pneumonia.

⍰ Scores of 4-6 → Class A → Low-risk → probability of progression <10%.
⍰ From 7-9 points → Class B → intermediate-risk → probability of progression of 10-40%.
⍰ From 10-13 points → Class C → High-risk → probability of progression > 50%

### Statistical methods

Frequencies and percentages were presented for the qualitative and mean variables and standard deviation for the quantitative variables based on the normality tests (P>0.05). The area under the model curve and the operational characteristics (sensitivity, specificity) were calculated for the best cut-off point. 95%confidence intervals (IC-95%) were presented. The data were processed in the SPSS statistical software version 20.

## RESULTS

### Participants

We found 120 patients who were admitted to hospitalization during the study period. Nevertheless, we included 51 patients to analyze.

### Descriptive data

The most frequent gender was male (66.7%; n=34). The mean age was 55.6 ± 15 years. The most frequent comorbidity was obesity (23.5%; n = 12). The median for LDH and absolute lymphocytes was 360 mg/dL (range: 208-1054) and 1100 lymphocytes/mm^3^ (range: 298-5671) respectively. The median Call Score was 6, with a range of 4 to 13. When they were divided into risk groups based on the CALL SCORE, the most frequent risk category was low risk (51%; n=26), followed by intermediate risk (33.3%; n = 17) and high risk (15.7%; n=8), respectively. See Table 1 for the general characteristics of the patients based on risk groups. A total of 8 patients died, representing 15.7%of the total. Eleven patients were admitted to the ICU (21.6%).

**Table 1.** General characteristics of the patients according to the Call Score risk group

|  |  | Low risk |  |  | Intermediate risk |  |  | High risk |  |  |
| --- | --- | --- | --- | --- | --- | --- | --- | --- | --- | --- |
|  |  | Frequency |  |  | Frequency |  |  | Frequency |  |  |
|  |  | (n) | Media | Median | (n) | Media | Median | (n) | Media | Median |
| Gender | Male | 18 |  |  | 10 |  |  | 6 |  |  |
|  | Female | 8 |  |  | 7 |  |  | 2 |  |  |
| Age |  |  | 48 |  |  | 60 |  |  | 71 |  |
| Arterial hypertension |  | 0 |  |  | 2 |  |  | 3 |  |  |
| Diabetes Mellitus |  | 0 |  |  | 2 |  |  | 5 |  |  |
| Obesity |  | 6 |  |  | 4 |  |  | 2 |  |  |
| Lactate dehydrogenase |  |  |  | 343 |  |  | 369 |  |  | 352 |
| Lymphocytes |  |  |  | 1289 |  |  | 739 |  |  | 621 |
| Death |  | 4 |  |  | 3 |  |  | 1 |  |  |
| Admission to the UCI |  | 4 |  |  | 3 |  |  | 4 |  |  |

### Main results

The CALL SCORE reported an area under the curve of 0.59 (IC-95%: 0.3 to 0.07), p=0.43 for predicting death (Figure 1). Nonetheless, for a cut-off point of 5.5, a sensitivity of 87%and a specificity of 65%were obtained. For the prediction of ICU admission, the area under the curve was 0.67 (IC-95%: 0.3 to 0.07), p=0.43 (Figure 2). The cut-off point of 5.5 presented a sensitivity of 82%and a specificity of 51%.

**Figure 1.**
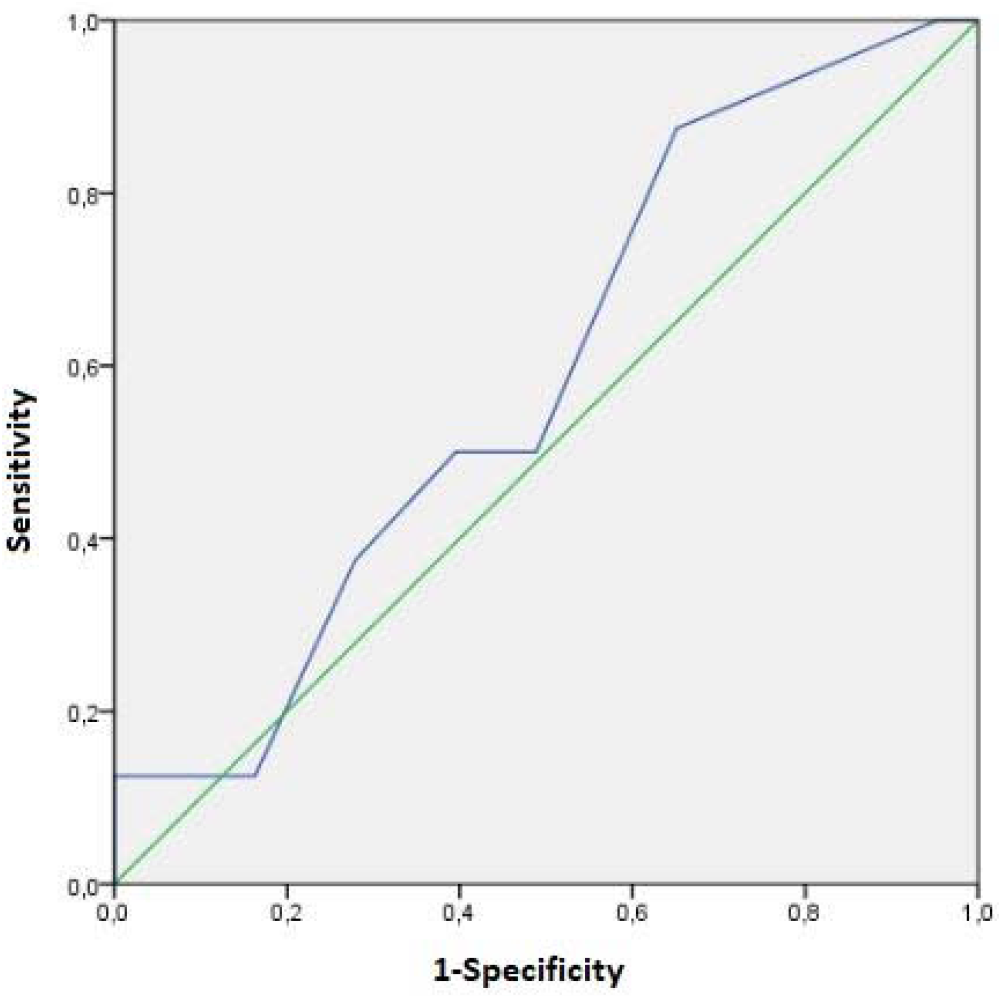
Clinical performance of CALL SCORE to predict mortality

**Figure 2.**
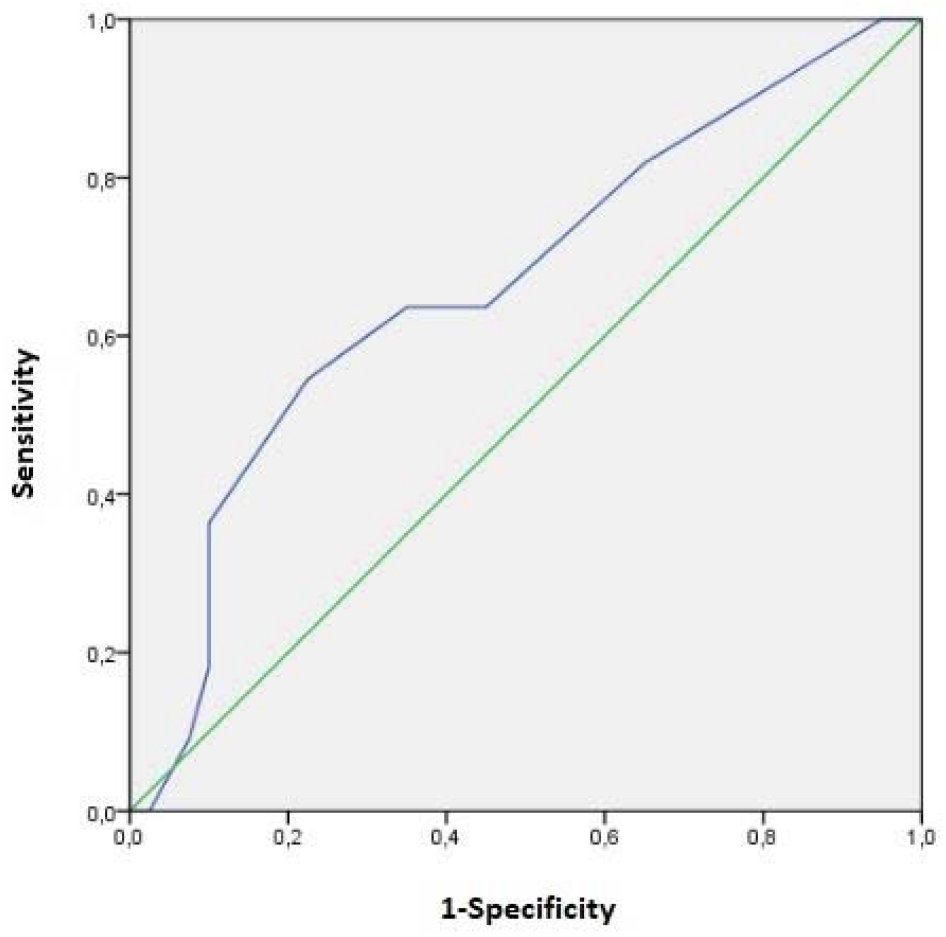
Clinical performance of CALL SCORE to predict admission to the UCI

## DISCUSSION

### Summary of the main results

The CALL SCORE showed a low performance for predicting mortality and admission to ICU in Peruvian patients. However, it presented a high sensitivity to predict admission to ICU and death with a cut-off point of 5.5.

### Contrast with the literature

These findings differ from the values reported in the derivation and validation study carried out in Chinese patients, with an area under the curve of 0.91 (IC95%: 0.86 to 0.94)[3]. Similarly, when the model was evaluated in the Italian population, the CALL score’s predictive power as a predictor of hospital mortality was good (AUC 0.768, IC95%: 0.705 to 0.823), differing from our result[8]. And in another study conducted in Pakistan, the call score was strongly associated with progression and mortality in patients with COVID-19[9]. The results show the importance of evaluating the validity of the clinical prediction rules developed in other countries for use in our population.

We observe that the performance differs from the two reported populations (China, Italy and Pakistan) and that its usefulness to evaluate mortality and severity in our country is limited.

Approximately 20%of COVID-19 patients are reported to develop severe respiratory illness, with an overall fatality rate of around 2.3%, and are secondary to COVID-19 complications, including acute respiratory distress syndrome. (ARDS), respiratory failure, liver injury, acute myocardial injury, acute kidney injury, septic shock, and even multiple organ failure[1]. In our study, 21.6%of patients were admitted to the ICU due to disease severity, and mortality was 15.7%. We did not evaluate the causes of admission to the ICU as it was not our study objective.

Patients with underlying comorbidities (hypertension, diabetes, pre-existing respiratory infection, cardiovascular disease, and cancer) are more likely to succumb and experience progression to the more severe forms of COVID-19. They are also at higher risk of developing complications[10]. In a document carried out by the Unit of Evidence and Deliberation for Decision Making of the University of Antioquia, they found that for mortality, severe illness and admission to the ICU, age over 60 years, as well as cardiovascular disease, arterial hypertension, and diabetes mellitus increase the risk[4]. We found that the most frequent comorbidity was obesity, followed by Diabetes Mellitus 2 and arterial hypertension, respectively. When classified by risk groups according to the CALL SCORE, these comorbidities were more frequent in the intermediate and high groups. Association measures were not performed because it was not the objective of the study. However, the predominance of these diseases can be observed in patients with a higher risk of severe disease, consistent with the literature.

As the pandemic has progressed and the disease is more widely known, it is found that various characteristics, including laboratory results, imaging studies, among others, are associated with a greater risk of hospitalization, death, or stay in intensive care units[3]. We found that the higher the risk group for the severe disease was, the more frequent lymphopenia was. However, we did not see the same behavior with elevated LDH.

The CALL SCORE’s low performance or clinical validity does not allow adequate prediction of mortality and admission to the ICU in our population. Apparently, with scores of 6, screening could be carried out due to the high sensitivity extended to patients not necessarily hospitalized.

We do not recommend using the CALL SCORE to predict mortality or admission to the ICU in Peruvian patients. It is probably useful as a screening test due to its high sensitivity in patients presenting to other services such as the emergency.

## Conclusions

The CALL SCORE presents a low clinical performance for predicting mortality and admission to the ICU in Peruvian patients. We do not recommend its use for the classification of patients requiring hospital management. The different clinical prediction rules used in our environment should be evaluated. The study should be replicated in other establishments and countries in South America.

## Data Availability

Dos de Mayo National Hospital Archives, Database, Lima-Peru.

http://hdosdemayo.gob.pe/portal/estadistica/boletin-epidemiologico/

https://drive.google.com/file/d/1kaLJwk4IFwYSIa28-4rRWe1WqPJg3QwS/view?usp=sharing

## AUTHORS CONTRIBUTION

All authors contributed to the conception of the project idea, protocol development, data collection, and writing and approval of the final handwritten.

